# Deuterium metabolic imaging of the human abdomen at clinical field strength

**DOI:** 10.1101/2024.09.10.24313302

**Authors:** Pascal Wodtke, Mary A McLean, Ines Horvat-Menih, Jonathan R Birchall, Maria J Zamora-Morales, Ashley Grimmer, Elizabeth Latimer, Marta Wylot, Rolf F Schulte, Ferdia A Gallagher

## Abstract

**Background:** The Warburg effect is a hallmark of cancer and is characterized by increased glucose consumption and lactate formation. Deuterium metabolic imaging (DMI) is an emerging non-invasive MRI method for probing this metabolic reprogramming in the field of neuroimaging. Here we show the feasibility of the technique for abdominal imaging using a routine 3 T MRI system, which has previously presented significant technical challenges.

**Purpose:** This study aimed to translate abdominal DMI to clinical field strength by optimizing the radiofrequency coil setup, the administered dose of deuterium (^2^H)-labelled glucose, and the data processing pipeline for quantitative characterization of DMI signals over time in the kidney and liver, establishing a basis for routine clinical studies in the future.

**Materials and Methods:** Five healthy volunteers were recruited and imaged on 2 or 3 occasions, with different ^2^H-glucose doses (totalling 13 DMI scan sessions). DMI was performed at 3 T using a flexible 20 x 30 cm^2^ ^2^H-tuned transmit-receive surface coil. We have defined three novel quantitative parameters as metrics of metabolism and compared these between doses and organs.

**Results:** The careful positioning of a dedicated surface coil minimized unwanted gastric signals while maintaining excellent hepatic and renal measurements. The timecourses derived from the liver and kidney were reproducible and comparable across different doses, with a trend towards lower quantitative measurements with decreasing dose. An increase in the ^2^H-water signal over time particularly in the liver, could be used as an indirect measure of metabolism.

**Conclusion:** DMI of the human abdomen is feasible using a routine MRI system and the metabolism measured in the kidney and liver can serve as a reference for future clinical studies. The ^2^H-glucose dose can be reduced from 0.75 to 0.25 g/kg to minimize gastric signal without substantially affecting the reliability of organ quantification.

## Introduction

Aerobic glycolysis, or the Warburg effect, occurs in a wide range of cancers and describes the increase in glucose breakdown which occurs even in the presence of abundant oxygen^1^. This effect is characterized by upregulated glucose consumption and the formation of downstream lactate that accumulates in the extracellular space, creating a hostile acidic extracellular tumor environment. A number of novel metabolic MRI methods have recently emerged to exploit this metabolism which have the potential for identifying more aggressive tumor subtypes and for assessing early metabolic response to treatments^2–5^.

In 2018, deuterium metabolic imaging (DMI) emerged as one of these non-invasive methods to assess glycolytic metabolism in the brain and cerebral tumors^6^. Unlike conventional proton (^1^H) MRI, deuterium (^2^H) is found at very low natural abundance (∼0.01%), and therefore deuterium spectra contain almost no background signal apart from the large peak arising from naturally-abundant, and partially-deuterated water (^2^H-water) signal, with some smaller deuterated peaks from adipose tissue. DMI exploits this low background signal through the administration of exogenous ^2^H-labelled molecules such as ^2^H-glucose ([6,6’-^2^H_2_]glucose)^7–11^. This labelled glucose is very safe^12^ and can be conveniently administered orally, thereby facilitating clinical use of the technique (Fig. 1a). Following absorption in the gastrointestinal tract, ^2^H-glucose metabolism can be followed spatially and temporally using deuterium magnetic resonance spectroscopic imaging (^2^H-MRSI)^6,9^.

**Figure 1.**
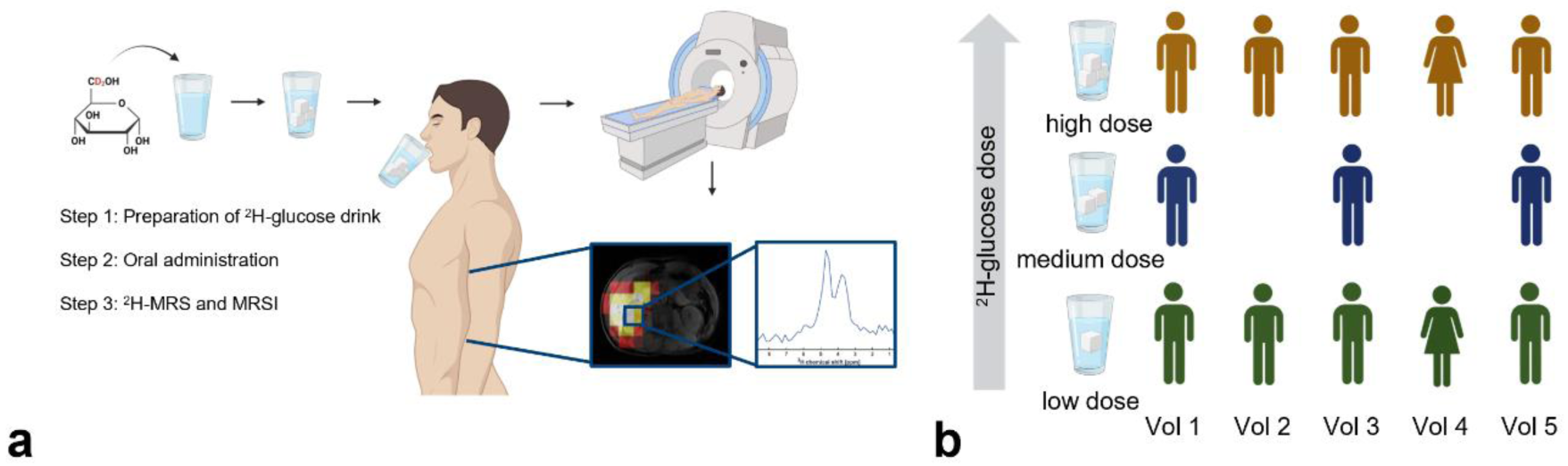
Study schema and experimental procedure. **(a)** DMI as a technique. A solution containing ^2^H-glucose is prepared and orally administered, followed by spectroscopic imaging. **(b)** n = 5 volunteers were imaged on two (n = 2) or three (n = 3) different days, totalling 13 imaging sessions. At each visit, the volunteers received one of three dose regimens of the ^2^H-glucose.

Despite having great potential for assessing glycolysis, DMI has faced several challenges in its translation into clinical field strength^7,13^. Whilst DMI has recently been translated to 3 T in the brain^9,14^, DMI outside of the brain suffers from additional challenges including: increased tissue inhomogeneity, motion, poor shimming, prominent gastric signal from the oral probe, and overlap between the naturally-abundant lipid peak and the tumor ^2^H-lactate signal, all of which reduce spectral quality. For these reasons, abdominal DMI has only been conducted at 7 T to date^8,10^.

In this study, we have addressed these challenges and conducted abdominal DMI in healthy volunteers at 3 T. The oral ^2^H-glucose dose has been varied to explore the optimal dose required for clinical imaging, to reduce both the cost and the gastric signal arising from the oral ingestion of the probe, without affecting the desired signal from the abdominal organs^15^. We demonstrate the feasibility of quantitative abdominal DMI of the liver and kidney at clinical field strength and present a robust pipeline for assessing ^2^H-glucose spectra for future clinical use including novel metrics of metabolism.

## Materials and Methods

### Study Design

The study was approved by a local research ethics committee (23/YH/0127) and all five recruited healthy volunteers (4 male, 1 female, age range = 20-30, Table 1) gave informed consent in writing. To assess the effect of dose reduction, the volunteers underwent up to three DMI studies on different days, receiving different doses of ^2^H-glucose (Cambridge Isotope Laboratories Inc., Tewksbury, MA), dissolved in water for injection (B. Braun, Melsungen, Germany), as shown in Table 1. The three dose regimens chosen for this study include the current standard dose given to volunteers and patients in previously published studies^9,11^ (“high”), as well as a dose reduction of one-third (“medium”) and two-thirds (“low”) of the standard dose. The “high” dose corresponds to 0.75 g/kg, with a maximum administration of 60 g. All five volunteers received the high and the low dose, while three volunteers also received the medium dose, totalling n = 13 DMI sessions. The volunteers were fasted for 6 h pre-scan. Prior to, and immediately after each exam, blood pressure, heart rate, blood oxygen saturation, and blood glucose level were recorded (Fig. S1).

**Table 1.** Demographics. Details about the individual volunteers and the dose received at each visit.

|  |  | <sup>2</sup>H-Glucose<br>[g] | Dose<br>[g/kg] | Dose label | Age<br>range<br>[a] | Sex | Height<br>[cm] | Weight<br>[kg] | BMI<br>[kg/m<sup>2</sup>] |
| --- | --- | --- | --- | --- | --- | --- | --- | --- | --- |
| **Vol 1** | Visit 1 | 60.00 | 0.69 | high | 26-30 | m | 192 | 87.0 | 23.6 |
|  | Visit 2 | 40.00 | 0.46 | medium |  |  |  | 87.0 | 23.6 |
|  | Visit 3 | 20.00 | 0.23 | low |  |  |  | 87.0 | 23.6 |
| **Vol 2** | Visit 1 | 20.00 | 0.19 | low | 26-30 | m | 189 | 108.0 | 30.2 |
|  | Visit 2 | 60.00 | 0.56 | high |  |  |  | 107.2 | 30.0 |
| **Vol 3** | Visit 1 | 40.00 | 0.50 | medium | 20-25 | m | 187 | 80.0 | 22.9 |
|  | Visit 2 | 60.00 | 0.75 | high |  |  |  | 80.0 | 22.9 |
|  | Visit 3 | 19.55 | 0.25 | low |  |  |  | 78.2 | 22.4 |
| **Vol 4** | Visit 1 | 40.88 | 0.75 | high | 26-30 | f | 164 | 54.5 | 20.3 |
|  | Visit 2 | 13.50 | 0.25 | low |  |  |  | 54.0 | 20.1 |
| **Vol 5** | Visit 1 | 33.50 | 0.50 | medium | 26-30 | m | 184 | 67.0 | 19.8 |
|  | Visit 2 | 50.18 | 0.75 | high |  |  |  | 66.9 | 19.8 |
|  | Visit 3 | 17.00 | 0.25 | low |  |  |  | 68.0 | 20.1 |

### ^2^H-MR Spectroscopy and Imaging

The volunteers were positioned supine in the MRI (3 T Premier, GE Healthcare, Chicago, IL) head-first. A flexible 20×30 cm^2^ ^2^H-transmit-receive surface coil (RAPID Biomedical, Rimpar, Germany) was wrapped around the right side of the abdomen, and fixed via Velcro straps, with the center of the coil positioned between the liver and kidney. An axial *T*_1_-weighted volume (LAVA-flex) was acquired for co-registration using the built-in body coil. Subsequently, a series of Hamming-filtered density-weighted ^2^H-MRSI (real matrix size = 10×10x10, 1678 interleaves, FOV = 40 cm, FA = 60°, TE = 0.7 ms, TR = 250 ms) were acquired for ≤90 min, interleaved between acquisitions, by the collection of unlocalized ^2^H spectra (TE = 0.7 ms, TR = 600 ms, FA = 90°, 64 averages).

### Data Processing and Analysis

Spectra were zero-filled by a factor of two in all three spatial dimensions and fitted automatically using a customised version of OXSA-AMARES^16,17^ with an SNR threshold of five. For display in Fig. 5, spatial interpolation by a factor of 12.8 in plane and a factor of 2 in the z-dimension was conducted. Voxels were manually assigned to the respective organs using an overlay of the summed ^2^H-signal and the axial anatomical LAVA-flex ^1^H localizer (FOV = 48 cm, FA = 12°, TEs = 1.1/2.2 ms, TR = 3.96 ms), as seen in Fig. 3. For liver, kidney, and duodenum, the prior knowledge of the fitting routine included three peaks: ^2^H-water, ^2^H-glucose, and ^2^H-lipid+lactate, as those commonly expected for DMI studies in the abdomen^10^. For the stomach, the prior knowledge was set to detect only one peak, assumed to be ^2^H-glucose in the absence of metabolism. Peak amplitudes, as fitted by OXSA-AMARES, were used to calculate the GGW and LLW (equation 1 and 2) ratio for every organ and timepoint using MATLAB (MathWorks, Natick, MA). The GGW and LLW ratio represent the respective normalization of ^2^H-glucose and ^2^H-lipid+lactate signals against the naturally abundant ^2^H-water peak to compare across scans as defined in equations 1 and 2:

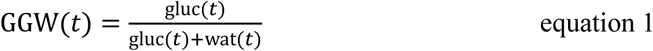

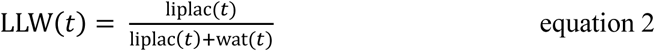

with gluc(t), liplac(t) and wat(t) representing the ^2^H-glucose, ^2^H-lipid+lactate (whose peaks overlap) and ^2^H-water signals at the respective timepoints. The GGW provides a normalized metric of ^2^H-glucose uptake, which is comparable across participants as a potentially useful clinical measurement. LLW is a similar comparative metric, which at baseline is dominated by the lipid signal and any increase over time corresponds to lactate formation in the organ of interest. For quantitative analysis of the timecourses, GGW_max_, GGW_AUC_, and GGW_mean plateau_ were calculated. GGW_max_ was determined by taking the maximum GGW value in each organ timecourse. For GGW_AUC_, the GGW value of each timepoint *t_n_* was multiplied by the time difference *t_n_-t_n-1_* between the previous timepoint in minutes (equation 3). The separate AUC(t) values were then summed until t = 74 min, which was a common timepoint across volunteers and exams, to make the GGW_AUC_ comparable across volunteers and across scans:

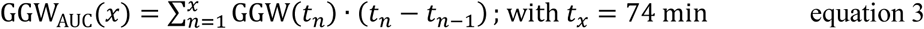

GGW_mean plateau_ was determined by taking the mean GGW of all timepoints after signal plateauing. The plateau time was determined to be t = 30 min for liver, and t = 40 min for kidney, broadly aligning with previously published studies^10^. For unlocalized spectroscopy, the GGW_mean plateau_ was replaced by GGW_mean_ over the whole timecourse, since the detection of signal build-up, and subsequent plateauing, was not possible for all scans, depending on the contribution of the stomach to the overall detected signal. For statistical analysis, Analysis of Variance (ANOVA) was performed in GraphPad Prism (GraphPad Software, Boston, MA), followed by Tukey’s Honestly Significant Difference (HSD) test as post-hoc analysis.

## Results

### Unlocalized Spectroscopy

Unlocalized spectra showed good SNR (> 5) for all three dose regimens, with the ^2^H-water (4.7 ppm) and ^2^H-glucose (3.7 ppm) peaks being clearly resolved at clinical field strength (Fig. 2a-c). Interestingly, the GGW ratio derived from these unlocalized spectra showed ^2^H-glucose build-up steadily over time in some volunteers, while appearing early in others (Fig. 2d). This is because, depending on the coil positioning, the stomach and duodenum were detectable in some cases, resulting in an early and high ^2^H-glucose signal in some cases. The low dose regimen exhibited a lower GGW across all metrics (Fig. 2e; GGW_max_ = 0.44 ± 0.03; GGW_mean_ = 0.35 ± 0.05; GGW_AUC_ = 24.1 ± 5.0; n = 4) compared to the medium (GGW_max_ = 0.55 ± 0.07, p = 0.0902; GGW_mean_ = 0.48 ± 0.05, p = 0.0193; GGW_AUC_ = 28.8 ± 2.7; p = 0.5097, n = 3) and the high doses (GGW_max_ = 0.57 ± 0.07, p = 0.0247; GGW_mean_ = 0.47 ± 0.05, p = 0.0148; GGW_AUC_ = 33.8 ± 6.4, p = 0.0560; n = 5). Statistically significant results are indicated in Fig. 2e. Notably, there was no significant difference in quantitative values between the medium and the high dose across all the metrics. The dynamics of the fitted ^2^H-glucose signal amplitude showed a signal build-up over 60 min in most cases (Fig. 2f). An increase in the ^2^H-water signal over time could be observed in all cases, ranging from 10-56% when comparing the last timepoint with the first in each case (Fig. 2g-h, Fig. S2).

**Figure 2.**
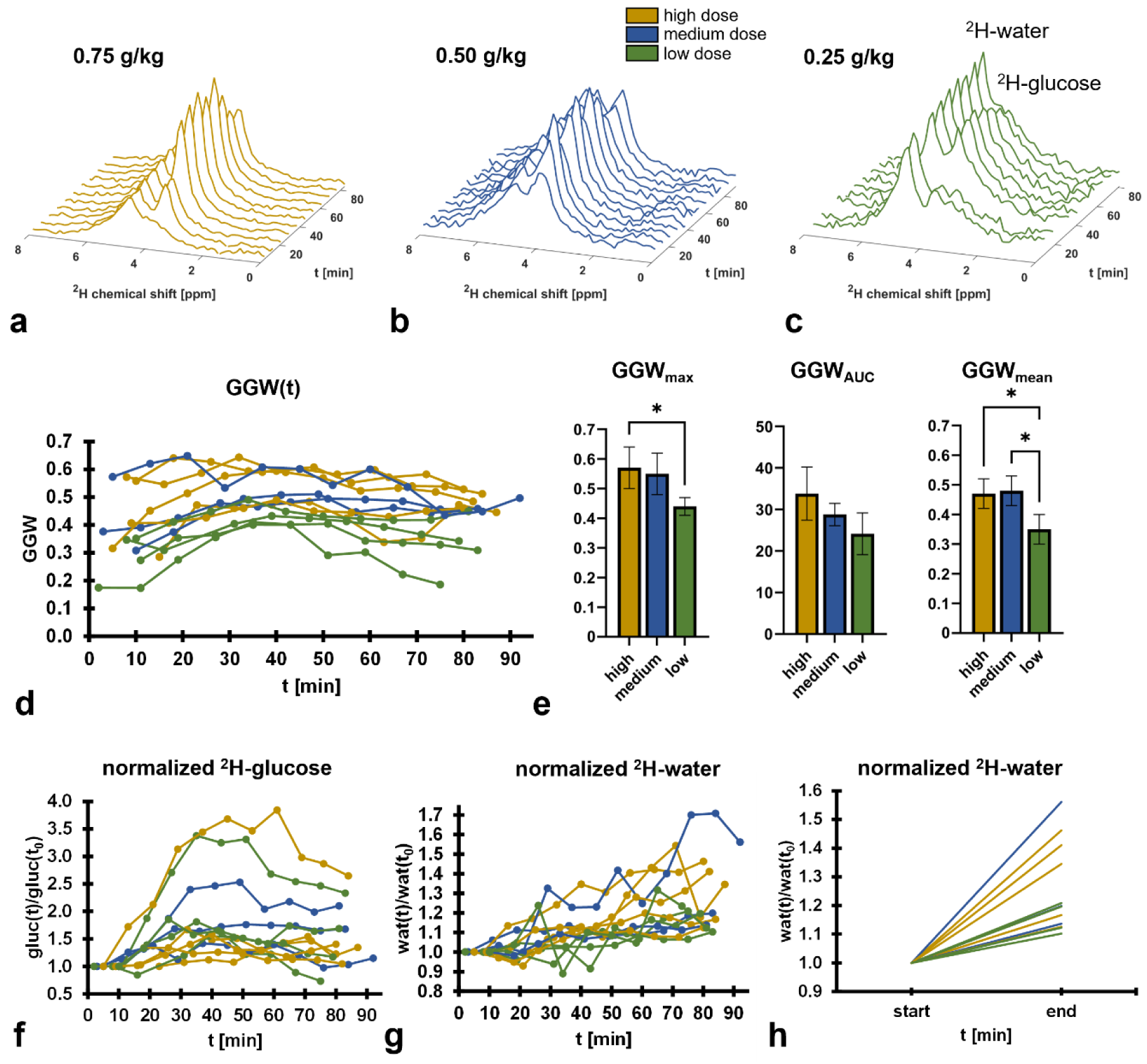
Dynamic unlocalized spectroscopy acquired from the healthy abdomen after oral administration of ^2^H-glucose. **(a-c)** Spectral timecourses demonstrating ^2^H-water and ^2^H-glucose peaks from a single volunteer (#3) after receiving the following ^2^H-glucose doses: (a) high; (b) medium; (c) low. **(d)** Timecourse of GGW (see methods) for each volunteer at each visit. **(e)** The three different quantitative measures we have introduced to evaluate ^2^H-glucose signal over time (see methods). **(f)** The total ^2^H-glucose signal initially increases with respect to the first timepoint in all volunteers and starts to decline after ∼50-60 minutes. **(g)** The ^2^H-water grows moderately over time in all volunteers, hinting suggesting the production of water through metabolism. **(h)** First (start) vs. last (end) timepoint of the data shown in (g), to demonstrate that all volunteers exhibit a growth in the ^2^H-water signal, thus suggesting a consistent effect.

### Imaging

Anatomically constrained ^2^H signal was identified in the liver and kidney on MRSI in all cases (n = 13; Fig. 3), and in the stomach and duodenum in most cases (n = 10 and 11 respectively). Only ^2^H-glucose could be detected in the stomach, while the duodenum exhibited a small but detectable ^2^H-water peak. The kidney exhibited a small lipid peak which was probably secondary to partial volume effects from the surrounding visceral fat.

**Figure 3.**
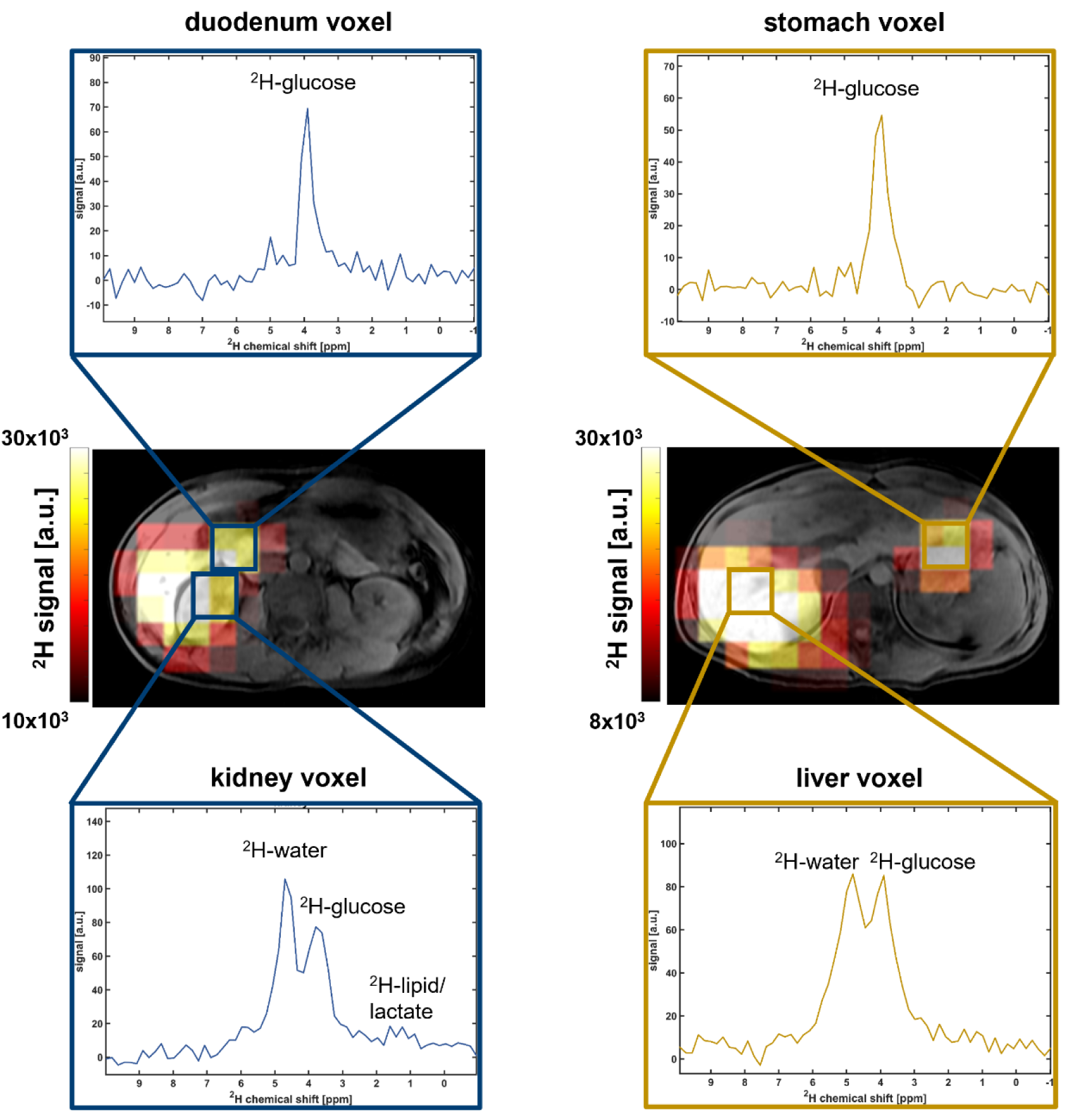
Representative images derived from spatially resolved spectroscopy. The summed ^2^H signal is shown as a colorscale image with single-voxel spectra from each organ (volunteer 5, t = 33 min after receiving the high (gold) and the medium (blue) doses. Stomach and duodenal ROIs are dominated by the ^2^H-glucose signal, whereas kidney and liver exhibit a prominent ^2^H-water peak as well as a small lipid peak in the kidney. The boxes in the images indicate non-interpolated voxel sizes.

The timecourses for the kidney (Fig. 4a) and the liver (Fig. 4b) showed a build-up in the GGW ratio, with the liver plateauing earlier at ∼30 min, compared to the kidney at ∼40 min. GGW signal in the duodenum remained high throughout the whole timecourse (Fig. 4c). The raw or non-normalized ^2^H-glucose signal was assessed in the stomach due to the low ^2^H-water signal, which showed a decrease over time in all cases as gastric emptying occurred (Fig. 4d).

**Figure 4.**
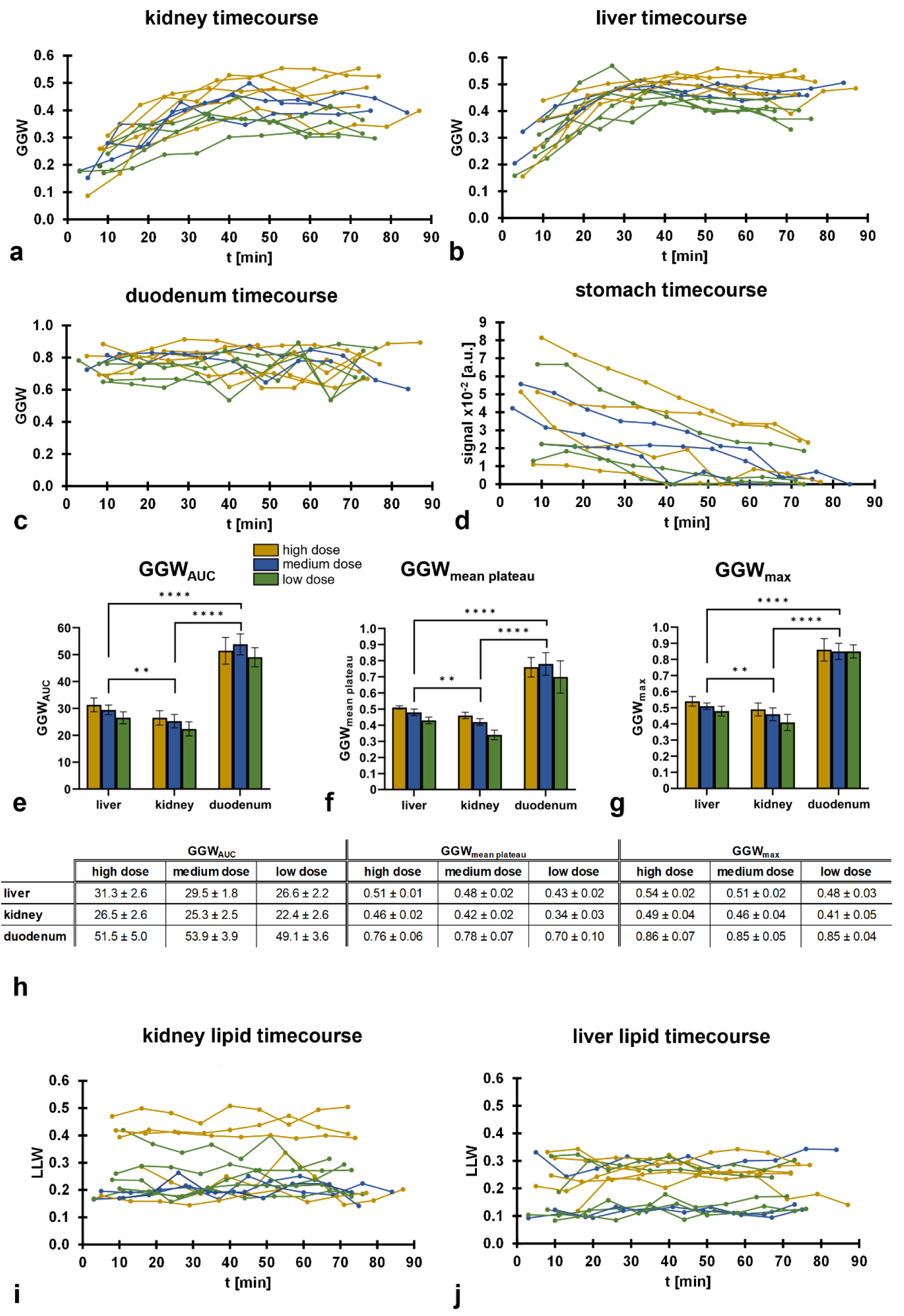
Quantitative evaluation of imaging data. **(a-c)** GGW timecourses for ^2^H-glucose signal in the kidney (a), liver (b) and duodenum (c). **(d)** ^2^H-glucose signal in the stomach over time shows a decrease likely to correspond to gastric emptying. **(e-g)** Quantitative measures of GGW_AUC_ (e), GGW_mean plateau_ (f) and GGW_max_ (g) exhibit dose dependence in the liver and kidney, whereas they remain elevated in the duodenum regardless of dose. **(h)** Summarized quantitative values displayed in (e-g). **(i,j)** ^2^H-lipid signal, as evaluated by the LLW parameter (see methods) over time. The kidney (i) shows a generally higher LLW than the liver (j), indicating a higher lipid content inside the imaged kidney voxels, which is likely to arise from the surrounding visceral fat.

### Comparison between the liver and kidney, and between doses

Across all three dose regimens and all three quantitative metrics, the liver exhibited a significantly higher ^2^H-glucose uptake than the kidney (Fig. 4e-h, p-values for GGW_max_/GGW_mean plateau_/GGW_AUC_: 0.0097/0.0032/0.0044). Furthermore, all three values increased with dose in both the liver and kidney. Notably, this trend was statistically significant between the low dose and the other two doses, but not between the high and the medium dose. Relevant p-values are shown in Figure S3. Example imaging timecourses from volunteer 5 for all three doses are displayed in Figure 5.

**Figure 5.**
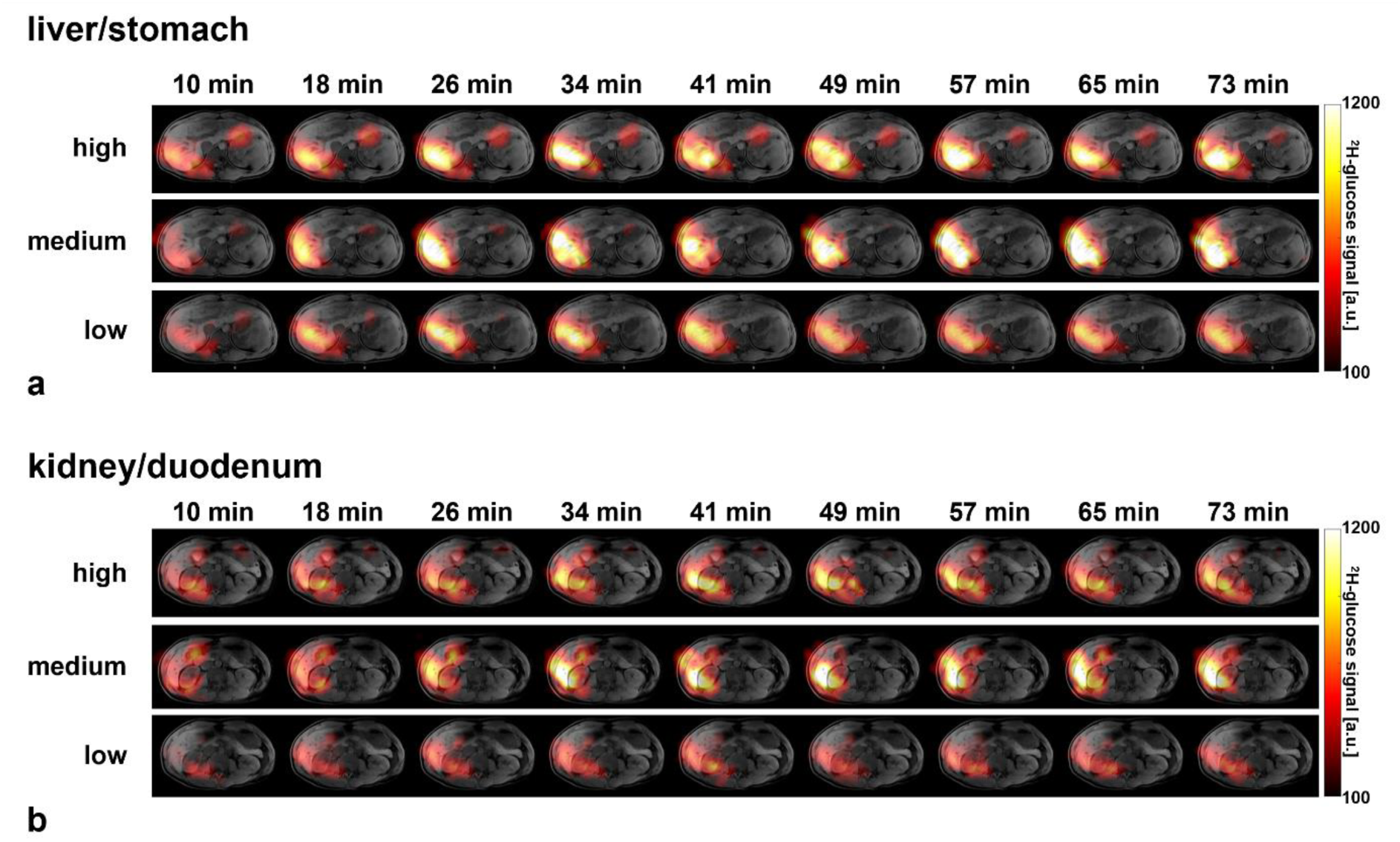
Imaging timecourses for ^2^H-glucose for all three dose regimens administered to volunteer 5. **(a)** A slice containing both the liver and stomach showing declining ^2^H-glucose signal in the stomach and increasing signal in the liver. **(b)** A slice containing both the kidney and the duodenum: the kidney shows a build-up of ^2^H-glucose, whereas the duodenum signal shows little variation in signal intensity over time. Timecourses of the ^2^H-water signal, as well as the summed ^2^H-signal can be viewed in the supplementary information (Fig. S4, S5).

### ^2^H-lipid content and ^2^H-lactate formation

The ^2^H-lactate signal appears at a similar chemical shift as the ^2^H-lipid signal (both at ∼1.3 ppm)^18^, and as lipid is particularly abundant in the abdomen^19,20^ this presents a challenge for the accurate quantification of ^2^H-lactate. Hence, it is more important to characterize the naturally abundant lipid signal on abdominal DMI, compared to the brain where the majority of previous DMI research has been undertaken.

A small ^2^H-lipid peak was identified in voxels overlying the kidney, and to a lesser extent the liver, which was likely due to partial volume effects from the visceral fat surrounding these organs (Fig. 3). The LLW ratio was calculated and the respective timecourses for both organs are displayed in Fig. 4i/j. The lipid content was higher in the kidney (mean across all volunteers and timepoints: 0.27 ± 0.10) compared to the liver (0.21 ± 0.08).

Lactate plays an important role in many pathological processes and therefore it is key to quantify the background ^2^H-lactate signal derived from normal organs after oral administration of ^2^H-glucose as a reference. Fig. 6a shows the normalized ^2^H-lipid+lactate signal for the three volunteers that were administered all three doses. A direct comparison of the last timepoint compared to the first for all volunteers can be seen in Fig. 6b-c. In the liver, the ^2^H-lipid+lactate signal increased in 12/13 examinations, with an average increase of 38.5 ± 24.2% showing that the healthy liver exhibits detectable ^2^H-lactate 60-80 min after administering oral ^2^H-glucose. In contrast, the ^2^H-lipid+lactate signal increased in only 8/13 examinations in the kidney, with an average increase of only 5.0 ± 17%, suggesting very little lactate formation in this timescale.

**Figure 6.**
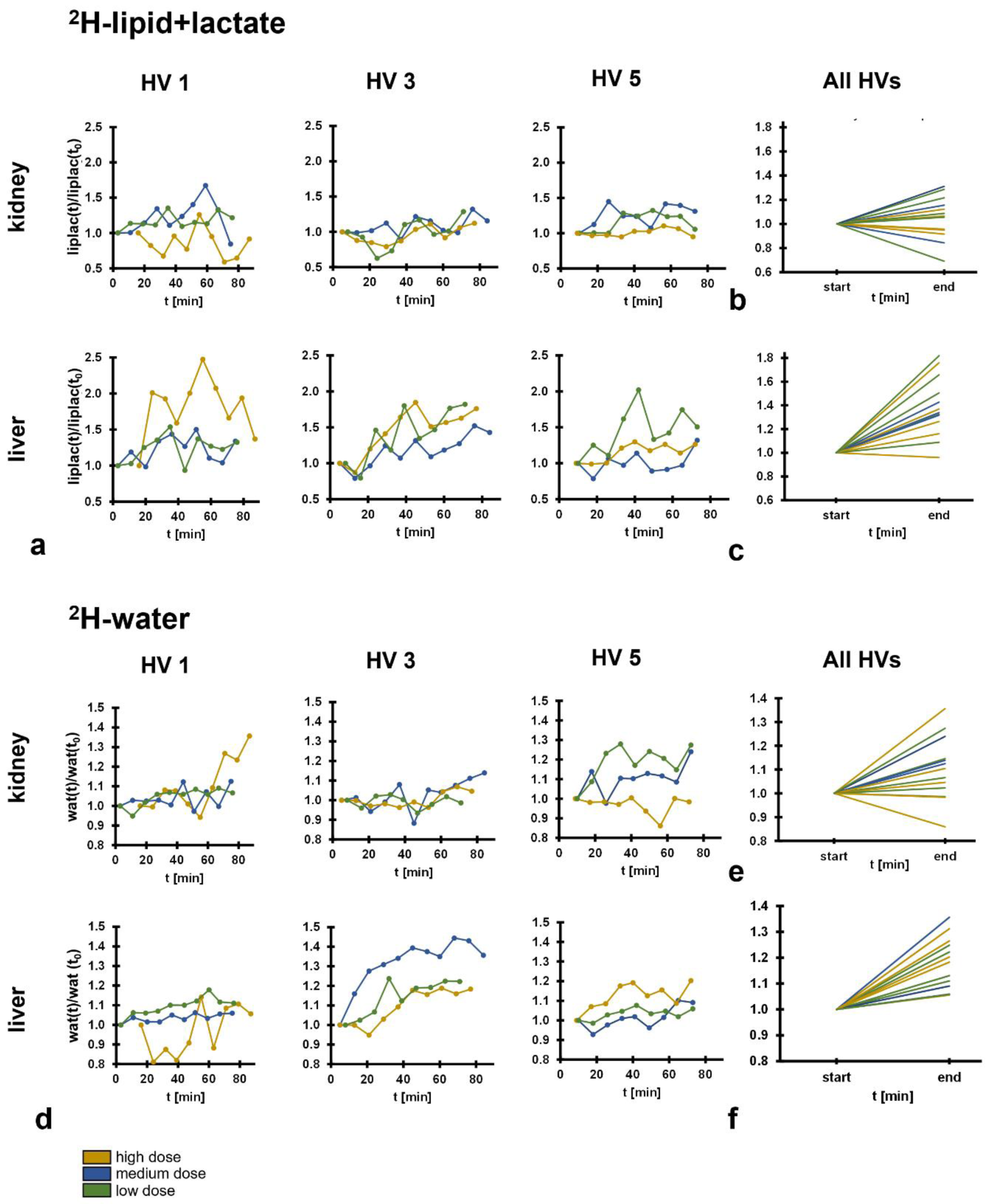
^2^H-water and ^2^H-lipid+lactate signal derived from individual volunteers over time, extracted from the imaging data. **(a)** ^2^H-lipid+lactate signal from the three volunteers that underwent all three imaging sessions. Unlike the kidney, the liver showed a trend towards an increasing signal over time, up to x2.5 the initial value. **(b,c)** Comparison of the first and last timepoints from all the volunteers reveals a trend towards increasing ^2^H-lipid+lactate signal in the liver (c), whereas the kidney (b) shows no trend across volunteers. The liver data suggests the hepatic formation of ^2^H-lactate from ^2^H-glucose in the timecourse of the experiment. **(d)** Comparative ^2^H-water signal of the same three volunteers. The liver shows a clear and consistent increase in ^2^H-water signal over time. **(e,f)** Comparison of the first and last timepoints from all the volunteers reveals a trend towards increasing ^2^H-water signal in both kidney (e) and liver (f) over time. In the case of the liver, the ^2^H-water signal increased in all volunteers.

### ^2^H-water signal increase as an indirect measure of metabolism

The unlocalized spectroscopy showed an increase in ^2^H-water signal over time (Fig. 2g-h), which has been reported in previous preclinical and clinical studies^9,10,21^. Spectroscopic imaging was subsequently used to localize this signal increase; Fig. 6d shows the normalized ^2^H-water signal for the three volunteers that were administered all three doses. A direct comparison of the last timepoint compared to the first for all volunteers can be seen in Figures 6e-f. In the kidney, the ^2^H-water signal increased over time in 10/13 examinations, with an average increase of 10.3 ± 12.8%. In the liver, the ^2^H-water signal increased in all 13 examinations, with an average increase of 17.6 ± 9.7%. Hence, the liver is likely driving the ^2^H-water signal increase in the unlocalized spectroscopy data. Indeed, the three examinations with the largest ^2^H-water signal increases in the unlocalized spectra were the same three examinations that exhibited the largest ^2^H-water signal increase in the liver imaging data. This observation further confirms the conclusion, that the liver is causing the dominant part of the ^2^H-water signal increase in the unlocalized spectra.

## Discussion

Deuterium metabolic imaging is an emerging method for probing tissue metabolism with potential applications in a wide range of diseases given the importance of glucose utilization in both health and disease. Here we translate the technique to the abdomen at clinical field strength, showing its feasibility for assessing hepatic and renal metabolism. We also optimize the dose of oral ^2^H-glucose for clinical use and present metrics to quantitatively evaluate its uptake and metabolism in tissues.

The signal from ^2^H-glucose was evaluated up to 90 min after oral administration and could be used to evaluate accumulation and absorption in the stomach and duodenum. Importantly, the use of a surface coil in combination with its positioning around the right side of the abdomen was used to minimize unwanted gastric signal when imaging the liver and kidney. The liver generally exhibited a higher ^2^H-glucose signal compared to the kidney, rapid plateauing of the signal, more reproducible time curves, and a lower lipid signal, therefore making it a promising organ to be assessed using DMI.

The increase in ^2^H-water signal over time is likely to arise from water derived from glucose metabolism^22,23^. The liver showed a higher ^2^H-glucose uptake than the kidney, as well as the formation of a small ^2^H-lipid+lactate signal, in keeping with increased hepatic metabolism. A confounder when analyzing DMI data is that the long timecourse for acquisition allows for metabolites from other organs to be washed into the organ under assessment so the measured signal may not be due to metabolism in the organ under investigation.

The doses of ^2^H-glucose used in DMI range from 60-75 g^8–11,24^, with 75 g being the standard dose for a clinical glucose tolerance test^25^. The blood glucose is transiently increased during these dose regimens, and we show that this could be avoided by lowering the dose to ∼20 g, allowing steady state metabolism to be probed without a substantial loss in signal (Fig. S1). This reduces the cost of DMI as well as the risk of unwanted gastric signal contaminating the quantification from other organs.

As an emerging technique, the best approaches for quantitative assessment and kinetic modelling of DMI are yet to be determined. The challenges are similar to other time-resolved metabolic imaging methods, such as those found in the analysis of positron emission tomography (PET)^26^ and hyperpolarized ^13^C-MRI data^27,28^. However, DMI also presents some problems unique to the technique and here we have proposed three novel quantitative biomarkers for assessment of this dynamic data: GGW_AUC_, GGW_mean plateau_, and GGW_max_. The results show that the hepatic and renal measurements are influenced by the administered dose but do not scale with it, suggesting a rate limiting element in delivery and metabolism. It is interesting to note that, despite the range of doses given across different volunteers, the duodenal measurements are remarkably constant over time and between individuals.

The increase in the ^2^H-lipid+lactate signal observed in the liver is likely to be from lactate formation as lipogenesis occurs on a slower timescale^29^. The lipid signal demonstrates a low SNR and has a broader linewidth compared to the ^2^H-water and ^2^H-glucose and may be overestimated while fitting with the automated approach here, which calculates a peak area.

One limitation of this study is the small cohort size; extending this study into larger cohorts and undertaking formal repeatability and reproducibility studies in a multisite setting will be important for future research in this field. Normalized plots have been referenced to the first timepoint of the same exam and this reference timepoint differed across examinations. It would be advantageous to reference the ^2^H-water and ^2^H-lipid signals against the naturally abundant measurements at baseline, but as the participants were repositioned after drinking in the upright position, the exact placement of the coil could change, potentially resulting in misregistration. Future studies could overcome this by maintaining the participant on the scanner with the coil in position and administering the ^2^H-glucose using a straw^8^, allowing the natural abundance scan to be directly compared to the post-administration images. Intravenous injection of ^2^H-glucose could also be considered in the future, bypassing gastrointestinal uptake, which would reduce the required dose further and lead to more rapid substrate delivery. However, this may result in a transient supraphysiological dose of glucose which could alter tissue metabolism.

## Conclusion

In conclusion, these data demonstrate the feasibility of quantitative DMI in the kidney and liver at clinical field strength, without signal interference by the stomach. Furthermore, we show that the administered dose can be significantly reduced, reducing the costs of a DMI scan and the otherwise dominant gastric signal, which are important considerations for future clinical use. Additionally, we introduced new methodology to assess glucose uptake and metabolism, which could be used to quantify tissue metabolism in disease settings.

## Data Availability

All data produced in the present study are available upon reasonable request to the authors.

## Acknowledgements

PW acknowledges support from the Gates Cambridge Trust (#OPP1144). MM acknowledges support from the Cambridge Experimental Cancer Medicine Centre. This research was further supported by Cancer Research UK (CRUK; C19212/A27150; C19212/A29082), CRUK Cambridge Centre, Wellcome Trust, NIHR Cambridge Biomedical Research Centre (BRC-1215-20014). The views expressed are those of the authors and not necessarily those of the NIHR or the Department of Health and Social Care. The authors also acknowledge support from the National Cancer Imaging Translational Accelerator (NCITA) and The Mark Foundation Institute for Integrative Cancer Medicine (MFICM) at the University of Cambridge.

