## Supplementary Information for "Deuterium metabolic imaging of the human abdomen at clinical field strength"

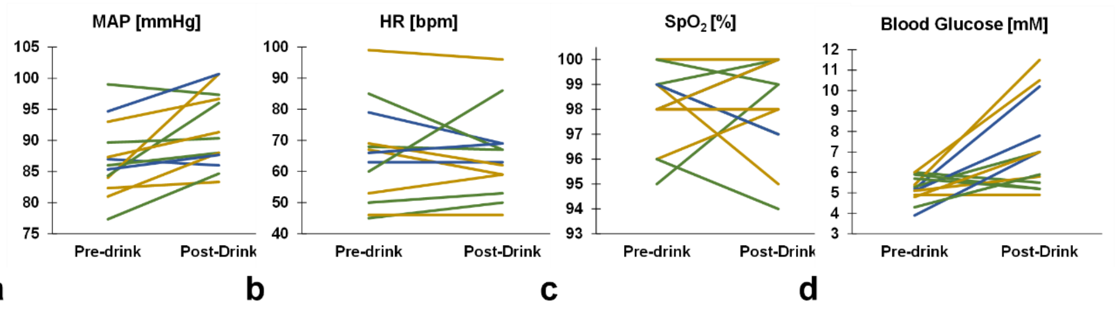


**Fig S1 | Physiological data acquired pre- and post-drink from the healthy volunteers.** MAP: mean arterial pressure; HR: heart rate; $SpO_2$: oxygen saturation.

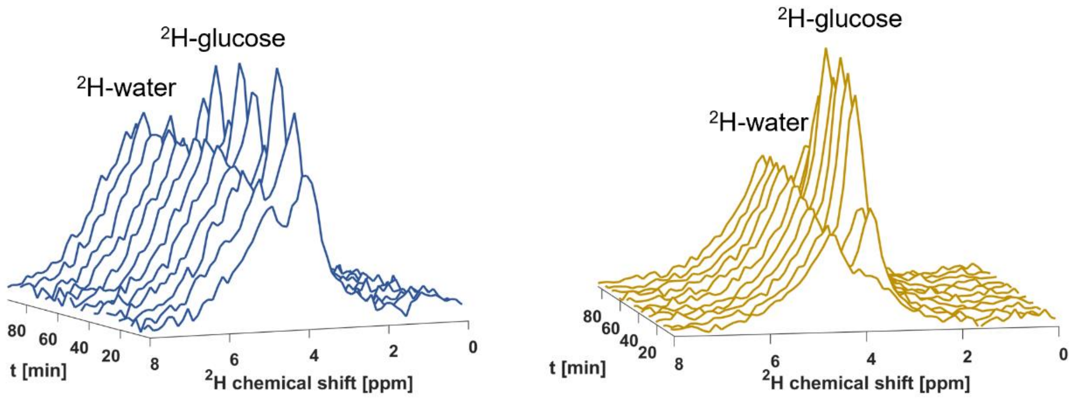


**Fig S2 | Representative spectra acquired from volunteer 3 after administration of medium (left) and high (right) dose, demonstrating a steady increase in the HDO signal over time.**

**$GGW_{max}$**

| | | p-value |
|---|---|---|
| Doses | high vs medium | 0.4667 (ns) |
| | high vs low | 0.0136 (*) |
| | medium vs low | 0.3827 (ns) |
| Organs | liver vs kidney | 0.0097 (**) |
| | liver vs duodenum | < 0.0001 (****) |
| | kidney vs duodenum | < 0.0001 (****) |

a

**$GGW_{mean\ plateau}$**

| | | p-value |
|---|---|---|
| Doses | high vs medium | 0.6904 (ns) |
| | high vs low | <0.0001 (****) |
| | medium vs low | 0.0053 (**) |
| Organs | liver vs kidney | 0.0032 (**) |
| | liver vs duodenum | < 0.0001 (****) |
| | kidney vs duodenum | < 0.0001 (****) |

b

**$GGW_{AUC}$**

| | | p-value |
|---|---|---|
| Doses | high vs medium | 0.9839 (ns) |
| | high vs low | 0.0084 (**) |
| | medium vs low | 0.0456 (*) |
| Organs | liver vs kidney | 0.0044 (**) |
| | liver vs duodenum | < 0.0001 (****) |
| | kidney vs duodenum | < 0.0001 (****) |

c

**Fig S3 | Statistical significance of MRSI data between doses and organs. (a) $GGW_{max}$; (b) $GGW_{mean\ plateau}$; (c) $GGW_{AUC}$.**

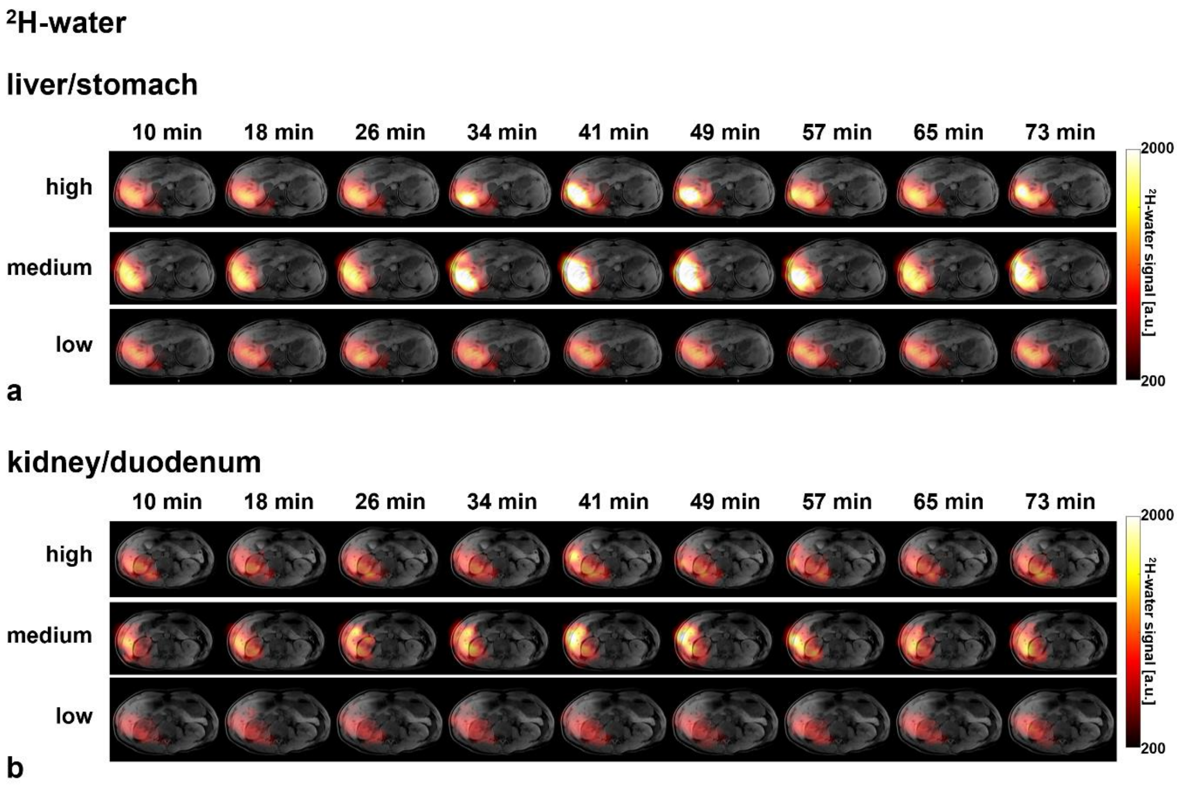


**Figure S4 | Imaging timecourses of the HDO signal for all three doses administered to volunteer 5.**

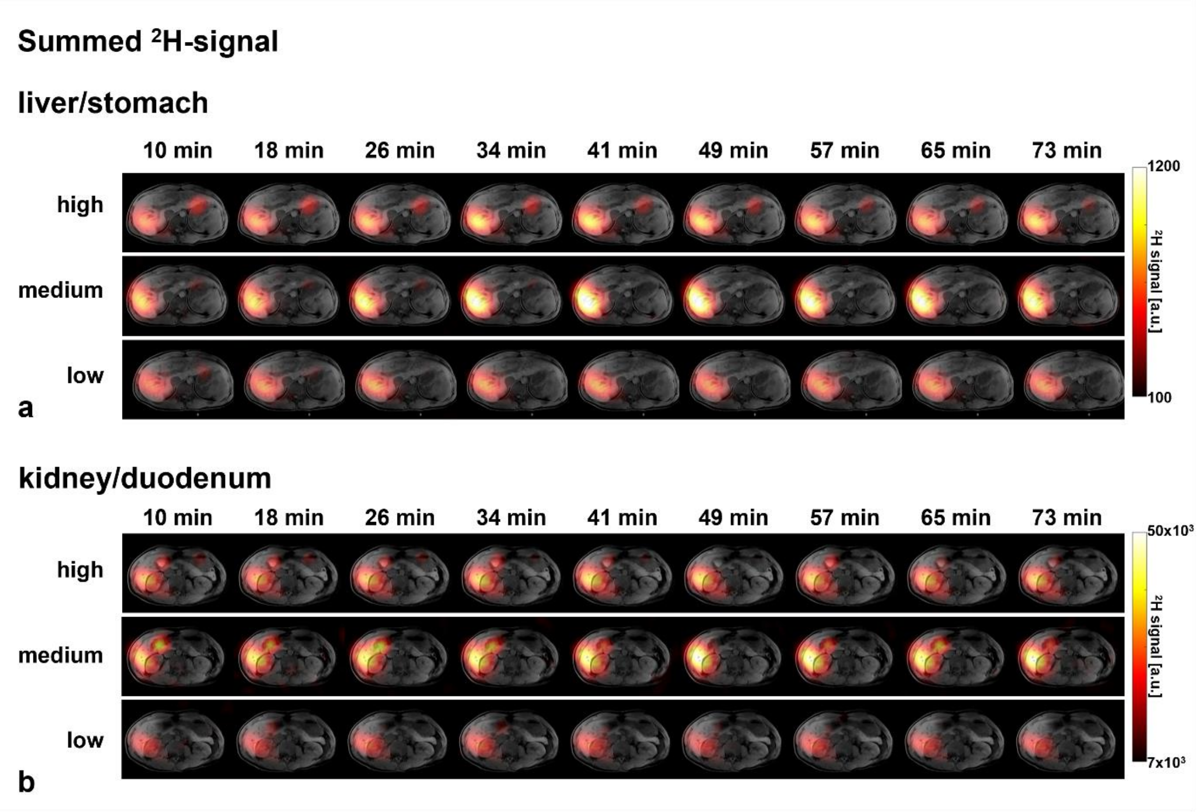


**Figure S5 | Imaging timecourses for the summed $^{2}$H-signal for all three doses administered to volunteer 5.**
